# Portability of an artificial intelligence model for self-harm detection across hospital settings

**DOI:** 10.1101/2025.07.10.25331160

**Authors:** Vlada Rozova, Katrina Witt, Mike Conway, Jo Robinson, Karin Verspoor

## Abstract

**Background:** Adequate self-harm surveillance is a key part of suicide prevention efforts. Our prior work has demonstrated the efficacy of an artificial intelligence model for detecting self-harm in emergency department triage notes. This model was developed based on data from a single hospital, raising the question about the model’s robustness to different contexts. Here, we aim to validate the model prospectively and externally to understand its portability across hospital settings.

**Methods:** Our self-harm classification model was developed and tested using triage notes from a large metropolitan hospital in Melbourne, Australia from 2012 to 2017. The model combined extensive text pre-processing with a Gradient Boosting classifier that used 644 selected features. In this study, we assessed the portability of both model components. We performed prospective validation using 329,655 triage notes from the same hospital collected over the following four years. For external validation, we used 316,877 triage notes from 2012 to 2021 from a regional hospital located 150km outside Melbourne.

**Results:** On the initial test set, the model achieved an area under the precision-recall curve (PR AUC) of 0.86, positive predictive value (PPV) of 0.81, and sensitivity of 0.80. Prospectively, the performance remained stable with PR AUC of 0.84, PPV of 0.76, and sensitivity of 0.76. Externally, the model showed a diminished ability to discern self-harm cases with an overall classification metric PR AUC of 0.77, PPV of 0.57, and sensitivity of 0.83. The text normalisation component of the model was equally effective across the datasets.

**Conclusions:** At the metropolitan hospital, the self-harm detection model is sufficiently performant for both epidemiological and potential clinical uses. At the regional hospital, the text normalisation pipeline is effective, but the machine learning classifier may need to be re-trained locally to produce more accurate results.

## Introduction

In Australia, emergency departments (ED) often serve as the primary, and sometimes sole, point of contact with healthcare services for patients following an episode of non-fatal self-harm. Self-harm is defined as any act of self-injury and/or self-poisoning irrespective of degree of suicidal intent and/or other motivations [1]. The accurate detection and monitoring of these events are crucial for public health initiatives, as they provide insight into self-harm prevalence, enable assessment of healthcare practices related to preventing repeated self-harm, and help estimate regional variation [2]. Moreover, given the strong association between self-harm – particularly when frequently repeated – and suicide [3], establishing robust self-harm surveillance systems can serve as an early warning mechanism for suicide and facilitate the evaluation of national suicide prevention strategies [4, 5].

However, relying solely on diagnostic coding within ED records often results in inadequate capture of self-harm incidents [6]. To improve the ascertainment of self-harm cases, nursing triage notes present a valuable, yet underutilised, source of information. These brief texts typically detail the reason for the ED visit, patient symptoms, physical appearance, and relevant medical history, potentially offering insights into the reasons and methods of self-harm.

We have previously developed an artificial intelligence (AI)-based model that uses natural language processing (NLP) and machine learning (ML) to detect self-harm from triage notes [7]. Our purpose was two-fold: a) to flag individual patient presentations that followed an episode of self-harm and b) to provide an estimate of the number of self-harm cases in the population. A near-perfect performance is essential for the clinical application. For epidemiological use, the positive predictive value (PPV) and sensitivity of the model are merely required to be balanced, since the overall trend is more important than accurately detecting individual cases.

The AI model was developed using data from the Royal Melbourne Hospital (RMH), one of the largest publicly funded hospitals in metropolitan Melbourne, Victoria, Australia. Triage notes from ED presentations between 2012 and 2017 were manually annotated by experts in suicide prevention as positive or negative for self-harm or suicidal ideation. The data was highly imbalanced with less than 2% of positive cases [8]. The model showed satisfactory performance at detecting self-harm cases, although some confusion with suicidal ideation cases was noted.

This study seeks to validate our model prospectively and to examine whether it can be used in a different setting. For prospective validation, we look at the next four years (2018 to 2021) of ED presentations at RMH. For external assessment, we obtained data from the Latrobe Regional Hospital (LRH). LRH is a public hospital serving a primary catchement area of over 300,000 persons residing in regional Victoria, Australia. It is located approximately 150km from the nearest large city, Melbourne. The catchment area is broadly representative in terms of age and gender but with a greater than average proportion of residents who identify as Aboriginal and/or Torres Strait Islander and a greater than average proportion of residents born in Australia.

## Background and Significance

Thorough validation of AI-based systems in healthcare is especially important given the potentially devastating consequences. In hospital settings, various factors can affect the performance of an AI-based system, ranging from differing patient demographics to site-specific protocols and data entry or management systems [9, 10]. Prospective and external validation studies are used extensively to ensure ML models generalise to unseen data [11-14]. Most validation studies are performed retrospectively to minimise the risks of premature deployment. The Epic Sepsis Model, a proprietary sepsis prediction model implemented by Epic in hundreds of US hospitals, has been an exception. The ease of integration within electronic health records allows researchers across multiple sites to validate the model locally and in real time [15]. Yet, studies report limited discrimination of sepsis onset [16, 17] and poor timeliness compared to traditional scoring algorithms [18].

Sociolinguistic variations across communities further contribute to dataset shift when analysing text-based documentation [19]. NLP models are additionally susceptible to performance issues due to variations in document structure and language [20], with extrinsic validation identified as critical [21]. Even with all other factors equal, different vocabulary and phrases used to describe conditions may severely affect the results. Approaches to validating NLP-based models include measuring the semantic similarity of corpora [22] and cross-checking models’ performance trained on different datasets [23]. At least some customisation is often required, either adjusting the text normalisation process [24, 25] or re-training the ML classifier [26].

As described in our previous work, nursing triage notes are often documented hastily and contain higher than typical levels of spelling mistakes. An example of a note could be, *“Taken 20×5mg diaz. O/A GSC 15, drowsy but tearfull. PHx dep[ression. Multiple superficial lacs on L) arm from 2/7 ago*.*”* Excessive use of ED-specific terminology, abbreviations, and slang is also characteristic of the notes. Normalising such text presents a challenging task. Our self-harm classification model consists of two components: a text normalisation pipeline and an ML classifier.

In the current work, we aim to evaluate the model’s performance prospectively and externally and to pinpoint the potential reasons for differences in performance. We aim to answer the following questions:

1. Was the population of patients similar (i.e., were observable variables like age, sex, and arrival method equally distributed)?
2. Was similar language used to record the information (i.e., is the text normalisation component equally effective)?
3. Were similar expressions used to describe self-harm (i.e., does the ML classifier achieve comparable performance)?

Both components of our model can be customised to new data. However, the text normalisation pipeline requires a human in the loop to review the results of spelling correction, and the ML classifier requires data to be expert-annotated for self-harm. As such, we explore whether tuning the first component without obtaining self-harm labels can improve the downstream task performance.

## Methods

### DATASET PREPARATION

Records of ED presentations at RMH and LRH were extracted from the respective hospital information systems over the ten years from 2012 to 2021 (Figure 1). The fields contained patient sex, age, time of arrival to ED, arrival method, and nursing triage note. We removed empty and duplicated rows and ED presentations of patients aged eight years or younger, as it is often difficult to determine the intent behind self-inflicted injuries in younger children. Throughout the paper, we refer to data used for model development, testing, and prospective and external validation as *development, test, prospective validation*, and *external validation* datasets.

**Figure 1.**
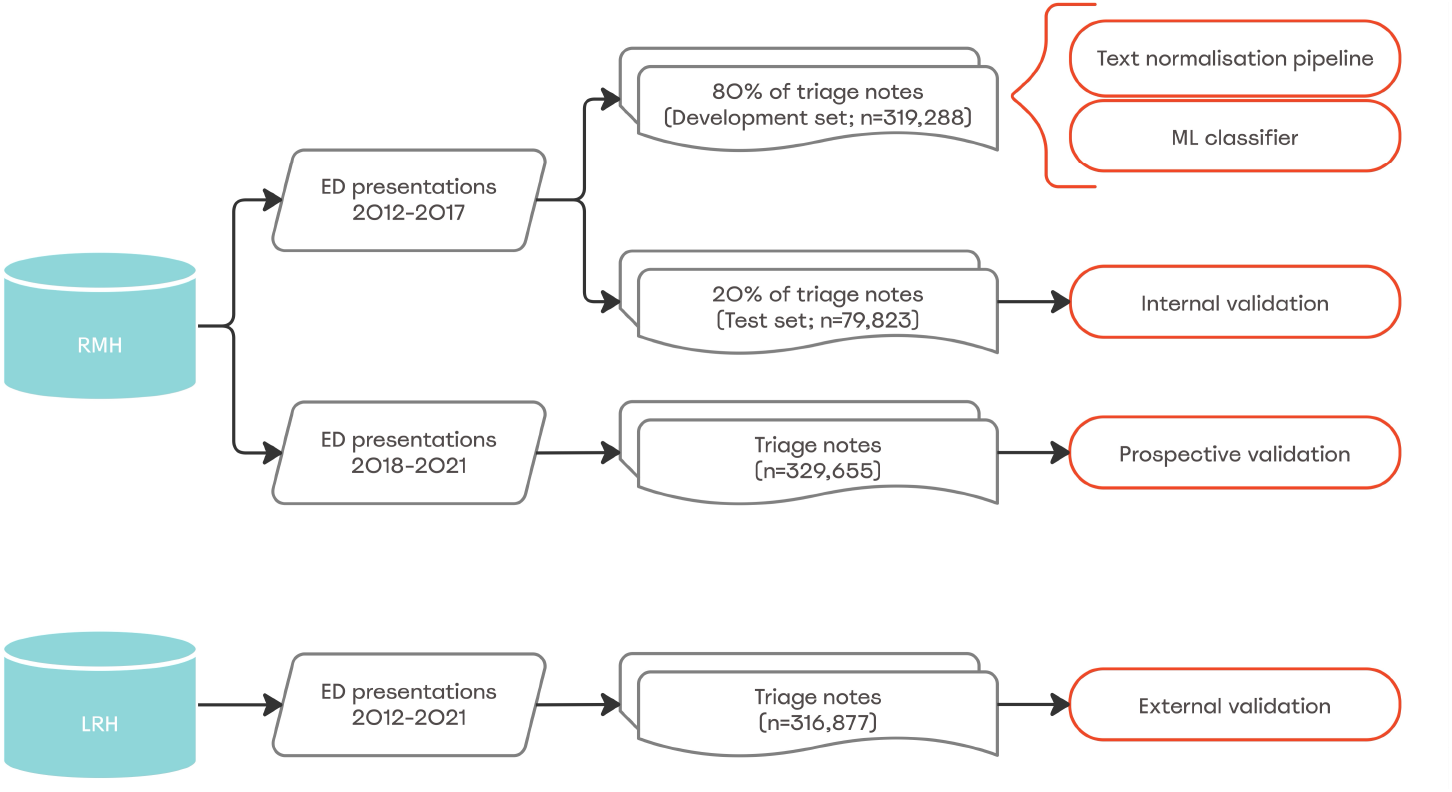
Triage notes used for the self-harm classification model development (80% of ED presentations at RMH in 2012-2017; *development* set), internal validation (20% of ED presentations at RMH in 2012-2017; *test* set), *prospective validation* (all ED presentations at RMH in 2018-2021) and *external validation* (all ED presentations at LRH in 2012-2021).

### DEVELOPMENT OF THE CLASSIFICATION MODEL

The development of the self-harm classification model is described in detail in [7]. Some modifications have been made since the publication of the paper to increase its robustness, which will be noted below. Broadly, the training phase consists of learning the vocabulary, learning the dictionary of misspellings, and fitting an ML classifier to predict self-harm. All components of the model were optimised for performance on the *development* set.

#### Learning the vocabulary

We iteratively constructed the vocabulary by leveraging several resources:

- Word lists from the SCOWL database [27] to include both general English words and Australian variants.
- A list of terms from the SNOMED CT-AU Australian ED reference set [28], for ED-specific terminology.
- Lists of terms from the SNOMED CT-AU medicinal and trade product reference sets [29, 30], for medication names.
- A comprehensive list of clinical abbreviations produced by the New South Wales Ministry of Health, Australia [31].
- A list of places in Victoria, Australia scraped from a local government website.
- A manually curated list of the most frequently occurring words in the *development* set that were not covered by any of the lists above (for example, names of local hospitals and their abbreviations, such as “RMH”).

In the final step, we filtered out words and abbreviations that did not appear in the *development* set. Given the relatively large number of triage notes (nearly 320,000), we reasoned that words not present in the *development* set are either very rare and do not carry important information or are not relevant to the ED context. The resulting vocabulary consisted of 20,115 words. Here, a word is a token that contains at least one letter: punctuation and numerical measurements are ignored, but “t2dm”, a common notation for type 2 diabetes mellitus, is considered to be a valid word.

#### Learning the dictionary of misspellings

For each out-of-vocabulary word in the *development* set, we attempted to find its correct spelling in the vocabulary. We built a dictionary of misspelled words and their correct spellings, yielding 43,909 pairs. Since finding candidates for a misspelled word is computationally expensive, we opted for a static approach and used the learned dictionary to correct spelling in unseen triage notes.

#### Selecting features and fitting a classifier

Once we had constructed the vocabulary and the dictionary of misspellings, we used them to normalise the notes in the *development* set. The *development* set was further split into five folds for cross-validation to determine the best approach to feature extraction (i.e., vectorisation), select the most important features, select the best-performing model, tune hyperparameters, calibrate the model and adjust the probability threshold. The final classifier was re-trained on the whole *development* set. It involved computing the term frequency-inverse document frequency (TF-IDF) matrix for 644 selected tokens and passing it into a LightGBM classifier for training and calibration.

### INFERENCE MODE

At prediction time, an unseen triage note undergoes the following steps: custom pre- processing, tokenisation, spelling correction, slang replacement for medication names, and removal of tokens that do not contain any letters. The result of the transformation below:

*“Taken 20×5mg diaz. O/A GSC 15, drowsy but tearfull. PHx dep[ression. Multiple superficial lacs on L) arm from 2/7 ago”* → *“taken x mg diazepam o/a gcs drowsy but tearful phx depression multiple superficial lacs on left arm from ago”*

A normalised triage note is then vectorised and passed into the pre-trained ML classifier to predict the probability of self-harm. The threshold is applied to convert the predicted probability to a class label.

### PERFORMANCE EVALUATION

To evaluate the performance of the self-harm classification model, we calculate the area under the precision-recall curve (PR AUC) and report PPV, sensitivity and specificity. To inspect changes in performance over time, we grouped triage notes by quarter (starting from January) based on the date of arrival to ED.

### LANGUAGE COMPARISON

Let us refer to the set of all valid tokens in the normalised notes from the *development* set as the *reference vocabulary*. We compared the language of triage notes against the *reference vocabulary* by calculating the proportion of shared tokens. We also report the total number of unique, valid tokens before and after text normalisation and how much it reduces dimensionality. Finally, we compared the feature distribution of each evaluation dataset with the *development set* by calculating their Jensen-Shannon divergence.

## Results

### MODEL EVALUATION

As described in the methods, the self-harm classification model was trained on the *development* set. We used the *test* set to evaluate the model internally, i.e., on unseen data from the same hospital and the same period of time. The model achieved a PR AUC of 0.85 during cross-validation on the *development* set and 0.86 on the *test* set. Table 1 reproduces these results along with performance on the *prospective* and *external validation* datasets.

**Table 1.** Overall performance of the self-harm classification model on the *development, test, prospective* and *external validation* datasets. Absolute differences with *test set* performance are shown in parentheses.

|  | Development set<br>(n=319,288) | Test set<br>(n=79,823) | Prospective<br>validation set<br>(n=329,655) | External validation<br>set (n= 316,877) |
| --- | --- | --- | --- | --- |
| PR AUC | 0.85 (+/- 0.01) | 0.86 | 0.84 (-0.02) | 0.77 (-0.09) |
| PPV | 0.81 | 0.81 | 0.76 (-0.05) | 0.57 (-0.24) |
| Sensitivity | 0.78 | 0.80 | 0.77 (-0.03) | 0.83 (+0.03) |
| Specificity | 1.0 | 1.0 | 1.0 (0.00) | 0.99 (-0.01) |

One can note a slightly lower performance when applying the model prospectively (PR AUC = 0.84, PPV=0.76, sensitivity=0.77). The probability threshold remains optimal, balancing PPV and sensitivity. However, when the model is applied at a different site, performance further drops with a nearly 20-point gap between PPV and sensitivity. This suggests the distribution of predicted probabilities is considerably skewed compared to the *test* set, potentially due to deeper-rooted differences in the underlying data (Figure S1). Adjusting the threshold (which would still require some annotated data) will not change the outcome — a lower PR AUC indicates that both PPV and sensitivity will be worse than on the *test* set.

Now, let us inspect the performance of the model over time. Figure 2 shows the changes in performance metrics when evaluated *prospectively* (RMH, 2018-2021) and *externally* (LRH, 2012-2021). We note a sudden decrease in performance at LRH starting from the second quarter of 2020, which coincides with the breakout of the COVID-19 pandemic in Australia. As a result, sensitivity drops to 0.68 by the end of 2021. Moreover, during the ten years, PPV remains considerably lower than sensitivity.

**Figure 2.**
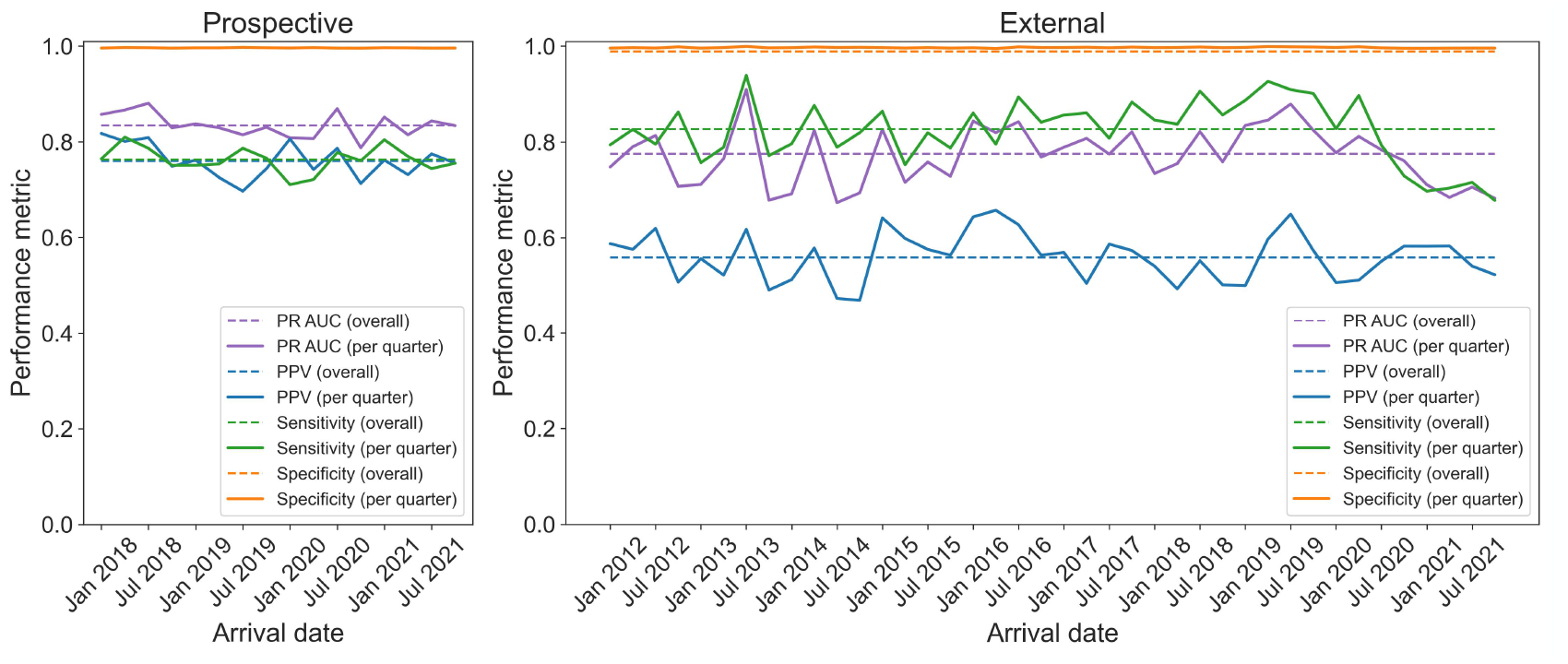
Performance of the self-harm classification model on the *prospective validation* (left) and *external validation* (right) datasets. Dashed lines indicate metrics calculated over the full sets of triage notes as reported in Table 2. Solid lines show the changes in metrics over time where triage notes were grouped by quarter based on the arrival date.

### COMPARISON OF THE TWO HOSPITALS

As noted in the introduction, multiple factors can lead to a decrease in performance. Table 2 summarises the key differences between the two hospitals provides an overview of the population statistics. One can easily see that the catchment areas and the number of ED presentations vary greatly; RMH also has a Mental Health team providing care in the ED. After removing presentations of children under the age of nine, RMH sees at least twice as many patients per year as LRH. Patient populations are similar in the distributions of age and sex; however, to get to the ED, more patients in regional areas rely on themselves / community than on ambulance services.

**Table 2.** Comparison of the two healthcare providers and patient population statistics from ED presentations between 2012 and 2021.

|  | RMH 2012-2021<br>(n=728,766) | LRH 2012-2021<br>(n=316,877) |
| --- | --- | --- |
| **Funding** | Public | Public |
| **Location** | Metropolitan | Regional |
| **Catchment area** | 137 sq.km | 42,000 sq.km |
| **Emergency Mental Health team** | Yes | No |
| **Number of ED presentations per year** | 72877 ($\pm 9517$) | 31688 ($\pm 4326$) |
| **Age** | 48 ($\pm 21$) | 47 ($\pm 23$) |
| **Sex** |  |  |
| Female | 48% | 52% |
| Male | 52% | 48% |
| Intersex | <1% | <1% |
| Unknown | <1% | <1% |
| **Arrival method** |  |  |
| Self / community | 23% | 47% |
| Road ambulance | 35% | 29% |
| Private ambulance | 1% | 2% |
| Air ambulance | <1% | <1% |
| Police | <1% | <1% |
| Undertaker | <1% | 0% |
| Other | 39% | 22% |

The overall proportion of self-harm cases over the ten years at RMH vs. LRH was 1.4% and 1.7%, respectively. The proportion of suicidal ideation cases was only slightly higher, 1.5% vs. 1.9%. The number of ED presentations per quarter at each hospital and the trends in self-harm and suicidal ideation cases over time are shown in Figure S2.

The typical length of triage notes indicates how much information is documented during ED encounters (Figure S3). Compared to the *development* set, triage notes at RMH have remained the same length. At LRH, triage notes, on average, were at least twice as long.

### THE EFFECTS OF TEXT NORMALISATION

To further explore where the differences in predicted probability distributions stem from, we evaluated the effects of the text normalisation pipeline. We compared the language of the *test, prospective* and *external validation* sets to the *reference vocabulary*. Before text normalisation, the number of unique tokens was the highest in the *prospective validation* set. In all three datasets, fewer than 50% of tokens were shared with the *reference vocabulary*. After text normalisation, the number of unique tokens dropped to approximately 10,000 in all three datasets. In *prospective* and *external validation* datasets, the proportion of tokens shared with the *reference vocabulary* increased to around 75%. A sudden increase in language similarity seen in the *external validation* data around April 2019 coincides with the roll-out of a new information management system at LRH. Text normalisation reduced the dimensionality of triage notes by about 50% in the *prospective validation* data, being slightly less effective on the *test* and *external validation* set. Taken together, these results do not reveal any dramatic differences in the underlying vocabulary that could explain the observed drop in performance (Figure S4).

### FEATURE SELECTION

To verify the portability of the ML classifier, we examined the distribution of 644 selected features used as classifier inputs after text normalisation had been applied. Figure 3 shows that while a sufficient number of selected tokens remained in the notes, their distribution varied. In the *prospective validation* set, the divergence increased sharply in early 2020. In the *external validation* set, the distribution of selected features differed considerably from the training data throughtout the entire 10-year period. These observations lead us to the conclusion that the differences must lie in the linguistic context, in *how* words were used to describe self-harm rather than *which* words were used. In other words, the relationships between the 644 tokens that the ML classifier has learned from the *development* set may not translate onto the *external validation* set; the model has overfit the patterns in the *development* set.

**Figure 3.**
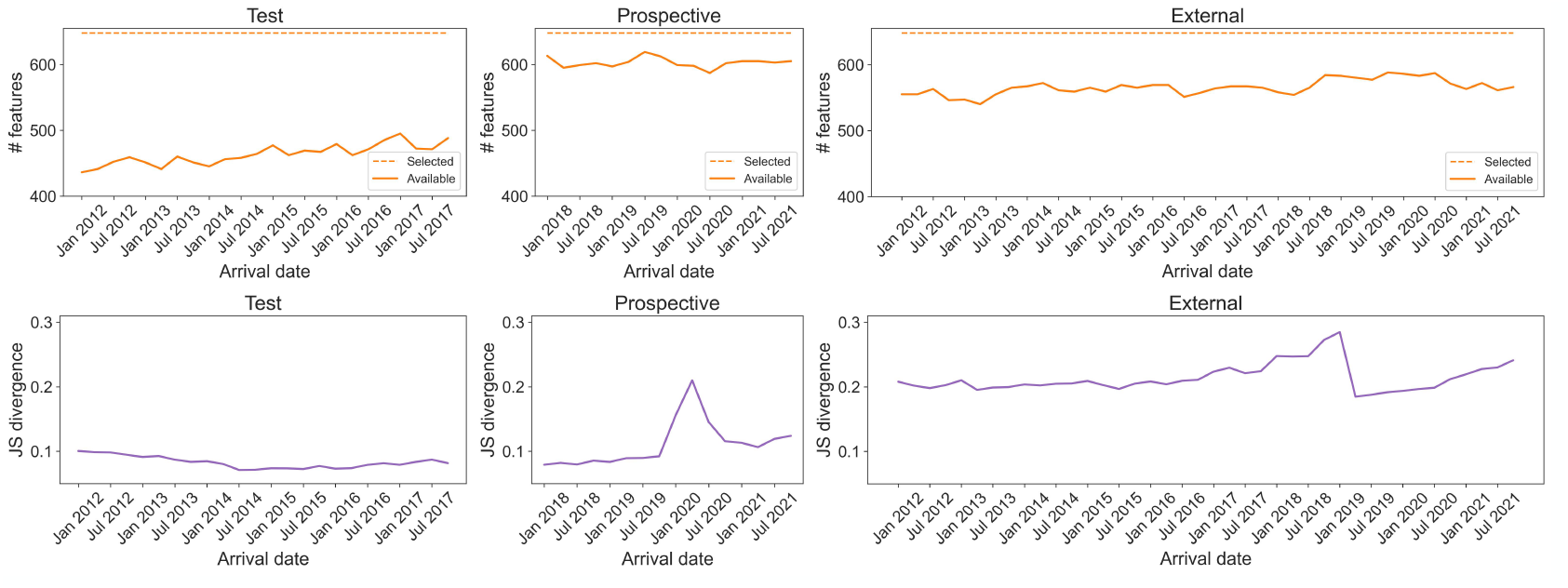
Features available after text normalisation in the *test* (left), *prospective validation* (centre) and *external validation* (right) datasets. Top row shows the number of available features (solid line) compared to the total 644 features used as input for the ML classifier (dashed line). Bottom row plots the Jensen-Shannon divergence with the *development* set feature distribution. Changes in all metrics are shown over time where triage notes were grouped by quarter based on the arrival date.

## Discussion

The challenges associated with the differences in the underlying training and test data distributions, known as dataset shift, have been known for decades. One of the earliest examples, as highlighted in [19], is a case of porting a non-NLP diagnostic ML model for acute abdominal pain from Leeds, UK to Copenhagen, Denmark. In that case, epidemiological and sociolinguistic differences between two contexts resulted in a drastic 27% decrease in overall accuracy. Various environmental, technological, and organisational factors may continuously impact model performance. During our study period, some observed factors included:

- A new information management system was introduced in April 2019 at LRH ED.
- Epic electronic health records system was introduced in January 2020 at RMH ED.
- A state of emergency was declared on March 16, 2020, in Victoria, Australia, in response to the COVID-19 global pandemic.

A plethora of frameworks exist guiding the development and implementation of AI models in healthcare. The majority advise validating the candidate model in a new healthcare setting or on data from a different period [32]. This is usually enough to see that a model fails to generalise, as continues to happen with the Epic Sepsis Model. However, a positive outcome of an external validation study does not guarantee the model’s universal generalisability [33].

Given the temporal nature of the factors affecting the data distribution, assessing the performance over time is especially important and can give insight into the actual model’s ability. Recurring validation locally may be preferred over a one-off external validation [34]. Moreover, focus should be put on evaluating aspects relevant to the desired application rather than attempting to prove all-round generalisability [9]. A model yielding similar PPV and sensitivity may be valuable in the epidemiological context, even if its overall discriminatory ability is not appropriate for clinical use.

## Conclusion

We show that a previously developed self-harm classification model remains useful in the original setting when validated prospectively. Balanced PPV and sensitivity suggested the model could be used to reliably track the proportion of self-harm cases in the population as an operatonal epidemiological tool embedded in public health workflows. Work is underway on deploying the AI-based system to facilitate self-harm monitoring at the RMH ED.

At the same time, the model was insufficiently performant to justify its implementation in the regional hospital. Our findings suggest that while the vocabulary of triage notes was similar, contextual differences not captured by the bag-of-words model resulted in decreased performance and, particularly, low PPV.

## Data Availability

All data produced in the present study are available upon reasonable request to the authors

## Ethical approval

The Self-Harm Monitoring System for Victoria has received ethical approval from the Melbourne Health Human Research Ethics Committee (HREC; 2017.342).

## Acknowledgements

We thank Associate Professor Jonathan Knott and Owen Conolly for facilitating data acquisision at the Royal Melbourne Hospital and the Latrobe Regional Hospital, Dr Michelle Lamblin as the program manager, Gowri Rajaram, Lu Zhang, and Hannah Richards for their assistance with manual data codng.

