## Supplemental materials for "Portability of an artificial intelligence model for self-harm detection across hospital settings"

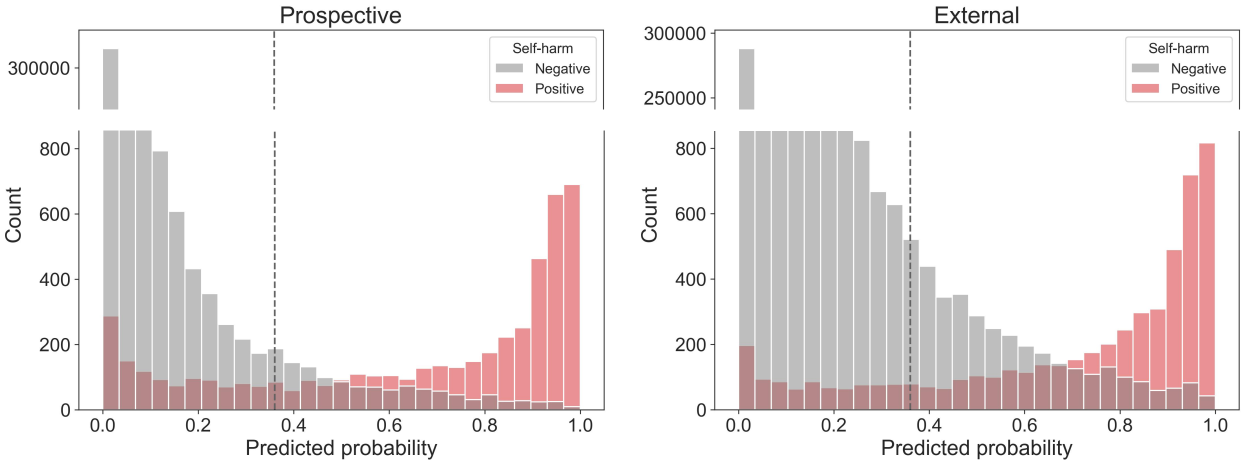


Figure S1. Predicted probabilities for the *prospective validation* (left) and *external validation* (right) datasets. Triage notes annotated as positive for self-harm are shown in red, controls are shown in grey. The dashed vertical line indicates the probability threshold applied to convert probabilities to class labels.

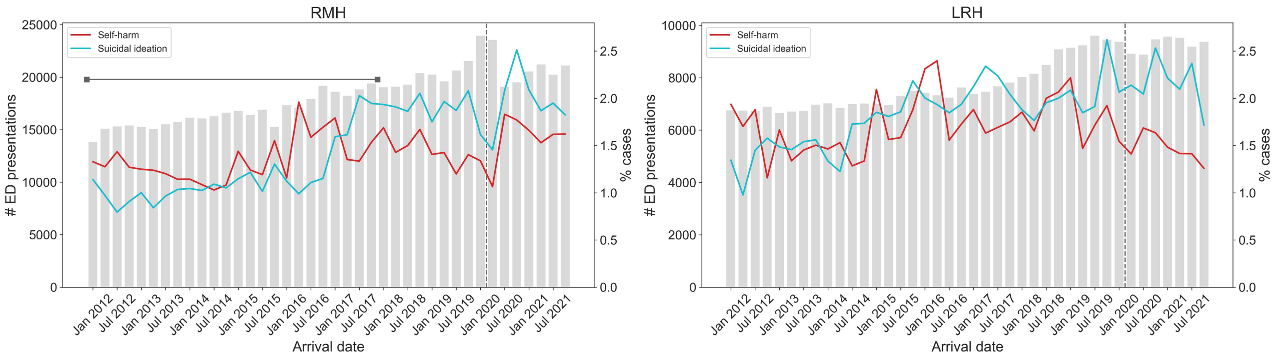


Figure S2. ED presentations grouped per quarter from 2012 to 2021 at RMH (left) and LRH (right). Grey bars show the total number of ED presentations. Changes in the proportion of self-harm and suicidal ideation cases are shown in red and yellow. The dashed vertical line indicates the start of the covid-19 pandemic in Australia. The first six years of RMH data were included in the *development* and *test* sets.

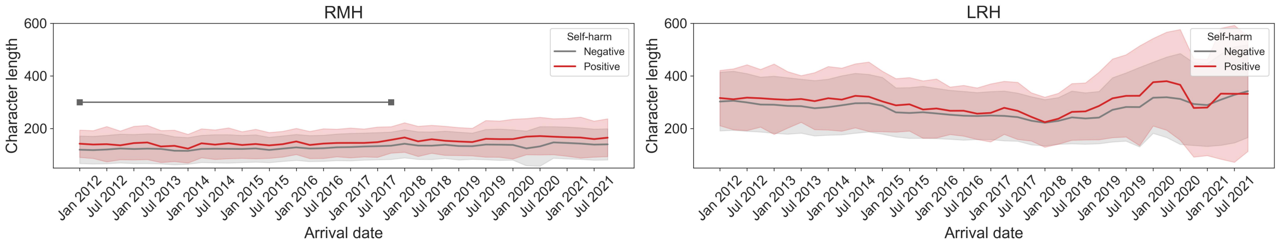


Figure S3. Character length of ED triage notes per quarter from 2012 to 2021 at RMH (left) and LRH (right). Triage notes annotated as positive for self-harm are shown in red, controls are shown in grey. Solid lines show the average length per quarter and the shaded area indicates the range as measured by standard deviation. The first six years of RMH data were included in the *development* and *test* sets.

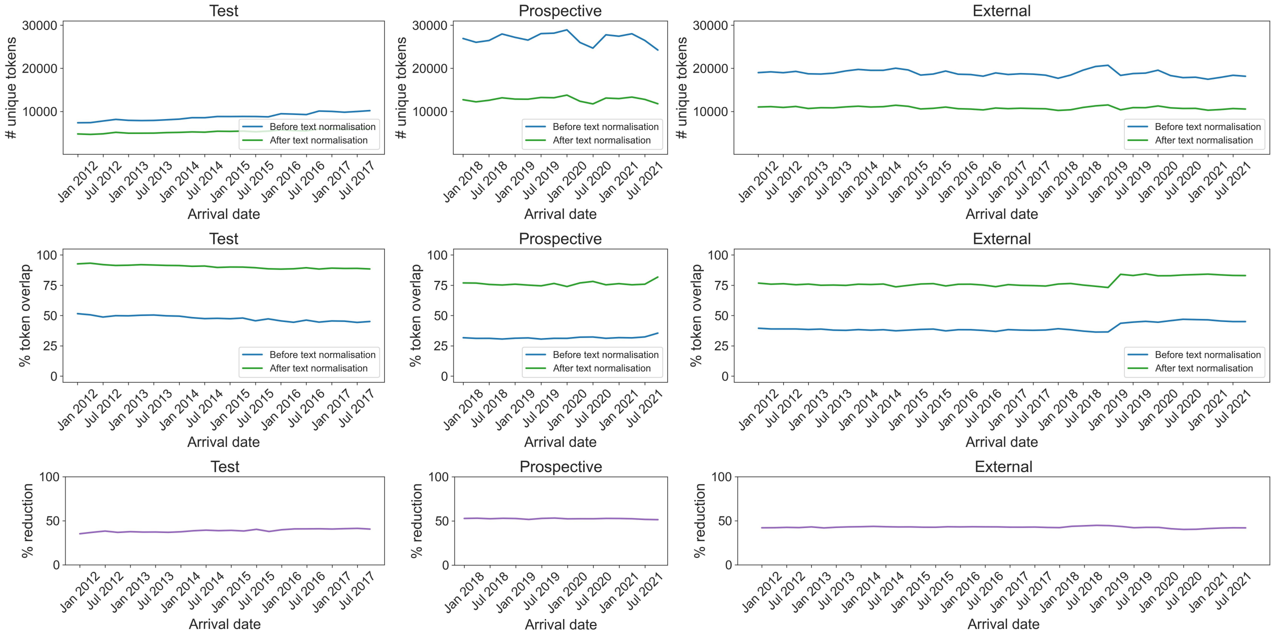


Figure S4. The effect of text normalisation pipeline on triage notes from the *test* (left), *prospective validation* (centre) and *external validation* (right) datasets. Top row shows the number of unique tokens in each dataset before and after text normalisation was applied. Middle row shows how many tokens overlapped with the *reference vocabulary* before and after text normalisation. Bottom row shows how much applying text normalisation reduced the dimensionality of data. Changes in all metrics are shown over time where triage notes were grouped by quarter based on the arrival date.
